# Machine learning-driven prediction of opioid and stimulant-related drug overdose fatalities: Analysis of the potential fourth wave

**DOI:** 10.64898/2025.12.09.25341941

**Authors:** Chisom Eze, Ryan Hansen, Marie Abate, Gordon Smith, Mohammad A. Al-Mamun

## Abstract

Between 2010 and 2021, fentanyl and stimulants co-involved deaths increased from 0.6% to 32.3% of all overdose deaths in the U.S. The Centers for Disease Control and Prevention monitors overdose deaths, but reports are delayed by about 4 -6 months. Therefore, advanced methods are needed for optimized trend monitoring and preparing the healthcare system. We developed and compared traditional and machine learning (ML)-based time series prediction models for forecasting opioid and stimulant-involved death rates. Forensic research data (2015 to 2023) built from the West Virginia (WV) Office of the Chief Medical Examiner data were used for this study. Decedents with any opioid or any stimulant-involved death were identified and placed into three cohorts ─ opioid-only, stimulant-only, and opioid and stimulant co-involved deaths. Monthly death rate per 100,000 was calculated for each cohort using total cases per month and West Virginia population data from Census Bureau. Autoregressive Integrated Moving Average (ARIMA), Random Forest (RF), and Extreme Gradient Boosting (XGBoost) variant models (differenced, non-differenced, and blended) models were trained on 80% of each cohort’s time-ordered data. An iterative forecasting of the 20% testing data was conducted. Model performance on the test prediction was evaluated using calculated metrics such as root mean square error (RMSE), R^2^, mean absolute error (MAE), and mean absolute percentage error (MAPE) values. Counts and percentages of cases per year were obtained for each cohort. Death rate and model predictions were represented in time series. Models’ performance for each cohort were compared using the performance metrics. 10,812 cases were identified from 2015 to 2023 with 4,295 involving opioid-only, 1,392 involving stimulant-only, and 4,175 co-involving an opioid and a stimulant. Stimulant-only and opioid and stimulant co-involved death rates had an upward trend with a peak in opioid and stimulant co-involved death in 2021. Although opioid-only death rate had a downward trend over time, the death rate peaked in 2020. The non-differenced XGBoost model outperformed for opioid-only (R^2^ = 0.92, RMSE = 0.12, MAE = 0.10, MAPE = 6.59%) and simulant-only (R^2^ = 0.91, RMSE = 0.07, MAE = 0.06, MAPE = 7.35%) death rate prediction. The blended XGBoost model had the best performance for opioid and stimulant co-involved death rate prediction (R^2^ = 0.78, RMSE = 0.31, MAE = 0.27, MAPE = 8.87%). Differenced XGBoost models outperformed other models for short term forecasting, while the non-different variants performed better for long-term predictions. Machine learning models, especially, the XGBoost variants outperformed other models for predicting opioid-only, stimulant-only, and opioid and stimulant co-involved death rates, respectively. The differenced models can be used for early death rate signal detection while the non-differenced XGBoost models can aid long-term forecasts for overdose death monitoring, planning and allocation of resources in health systems.

**Author summary:** The United States has been faced with the problem of opioid abuse and overdose death for several decades. Currently, there is a rise in drug overdose deaths co-involving an opioid and a stimulant. Although the CDC monitors and produces a provisional overdose death count, this report is often delayed by 4-6 months. There is a need to develop a high accuracy predictive tool that can yield reliable forecasts of these overdose deaths that can be used to guide policy decisions and avoid the delay. Here we developed and compared machine learning (Extreme gradient boosting (XGBoost) and Random Forest) to traditional statistical (ARIMA) forecasting models for predicting overdose death rates involving an opioid, a stimulant, or both. We found that the XGBoost models performed better than ARIMA and Random Forest for making the predictions. Our study provides a tool that can be used to predict future overdose deaths and provide information to prepare health systems and communities to better respond to overdose deaths and develop policies targeting drug, especially opioid and stimulant overdose prevention.

## Introduction

Synthetic opioid abuse, especially involving illegally manufactured fentanyl (IMF), became prominent in the United States in 2013 (1). More recently, synthetic opioid-involved fatality rates increased by 1,040% from 2013 to 2019, and the psychostimulant-involved death rates increased by 317% (1). Between 2010 and 2021, fentanyl and stimulant-involved fatalities rose nationally from 0.6% to 32.3% of all overdose deaths (2). In 2021, in West Virginia (WV), 76% of all drug overdose deaths involved fentanyl and fentanyl analogs, an 18% increase from 2017, with methamphetamine-involved deaths accounting for 52.2% of all drug overdose deaths (3).

Faced with a rising trend in opioid and stimulant co-involved fatality rates, the importance of proactive surveillance for early warning and hotspot detection, justifying funding decisions, and preparing the healthcare system cannot be overemphasized. The Centers for Disease Control and Prevention (CDC) has focused on trend monitoring, identifying severely affected areas, supporting state and local authorities and healthcare systems, and increasing public awareness to encourage safer individual choices (4). CDC provides Provisional Drug Overdose Death Counts based on reported and predicted death counts, but with a 4 – 6 months lag in reporting (5). Advanced methods are needed to produce overdose drug death estimates for early detection of misuse trends, evaluating intervention success, informing public awareness, preparing the healthcare system for emergencies and addiction services, and evidence-based policy making.

Generally, forecasting time series incorporates trends, seasonality, and cyclic patterns from past data collected over time to predict future values of a given outcome. Statistical methods such as Autoregressive Integrated Moving Average (ARIMA) have been widely used for time series analysis. Using proxy data from sources including the National Syndromic Surveillance Program and the National Forensic Laboratory Information System, Sumner et al. compared the weekly U.S. opioid overdose death count predictions from a least absolute shrinkage and selection operator (LASSO) regression model to a seasonal ARIMA (SARIMA) model (6). While the LASSO model made predictions with an error of 1.0% for the first year and −1.1% for the second year, the SARIMA model overestimated the weekly death counts by 32.8% the first year, then improved in the second year with a 4.1% error (6). Another study fitted an ARIMA model for estimates of U.S. overdose deaths from February 2016 to March 2020, then used the model to generate forecasts for the remaining 43 weeks in the latter year (7). Predictions by their model only differed by 12% from actual overdose deaths within that timeframe. Machine learning (ML) techniques such as Random Forest (RF) have been repurposed for forecasting drug overdose fatalities. Allen et al. outlined a practical and implementation-based framework for evaluating two different predictive models, Gaussian Process (GP) and RF, for predicting drug overdose death counts in Rhode Island (8). Relative to the GP model, RF predicted 10.2 – 36.4% of overdose deaths at different state-wide implementation capacities (8). With advances in predictive analytics, many newer and existing methods, including Extreme Gradient Boosting (XGBoost), long short-term memory, can be repurposed and evaluated for drug overdose death forecasting. These methods have the potential for developing even more accurate and precise models to optimize forecasts that can be used for surveillance, developing, and evaluating interventions, and allocating resources.

This work aimed to develop and compare the predictive performances of ML (RF and XGBoost) to a standard statistical (ARIMA) model for forecasting monthly fatality rates at the state level for opioid-only, stimulant-only, and opioid and stimulant co-involved deaths. The best performing model will be determined to provide reference information for opioid and stimulant-associated deaths.

## Results

### Population Characteristics

From 2015 to 2023, 4295 total opioid-only deaths occurred in WV (Table 1), with most deaths in 2016 (13.81%). A total of 1392 stimulant-only deaths were recorded within the same timeframe. Of the 1392 deaths, 15.45% occurred in 2019. A total of 4175 opioid and stimulant co-involved deaths were recorded from 2015 to 2023, with 18.28% of the deaths occurring in 2021.

**Table 1:** Number of decedents per year for opioid-only, stimulant-only, and opioid and stimulant co-involved fatalities.

| Year | Total Deaths per year | Opioid-only<br>N = 4295 | Stimulant-only<br>N = 1392 | Opioid and Stimulant<br>N = 4175 |
| --- | --- | --- | --- | --- |
|  | 10,812 |  | N (%) |  |
| 2015 | 746 | 489 (11.39) | 50 (3.59) | 117 (2.80) |
| 2016 | 1057 | 593 (13.81) | 100 (7.18) | 202 (4.84) |
| 2017 | 1150 | 575 (13.39) | 124 (8.91) | 326 (7.81) |
| 2018 | 1122 | 438 (10.20) | 171 (12.28) | 374 (8.96) |
| 2019 | 1046 | 351 (8.17) | 215 (15.45) | 362 (8.67) |
| 2020 | 1452 | 570 (13.27) | 175 (12.57) | 593 (14.20) |
| 2021 | 1555 | 502 (11.69) | 211 (15.16) | 763 (18.28) |
| 2022 | 1417 | 419 (9.76) | 167 (12.00) | 758 (18.16) |
| 2023 | 1480 | 358 (8.34) | 179 (12.86) | 680 (16.29) |

### Time Series Plots for Opioid, Stimulant, and Opioid and Stimulant Death Rates

The imputed monthly opioid-only death rate per 100,000 in WV had a fluctuating increasing and decreasing trend across the years studied (Supplementary file, Figure S1a), showing non-stationarity, which implies that the death rate changed over time. The opioid-only death rates showed an upward spike (4.92 per 100,000) in 2020 which coincides with the beginning of the COVID-19 pandemic, then dipped sharply before going through a gradual decline. However, there was no obvious seasonality observed in the time series plot. Imputing the spike in 2020, reduced the value’s deviation from the series’ mean. Stimulant-only death rate depicted an obvious upward trend from 2015 to 2019 (Supplementary file, Figure S1b). The trajectory was inconsistent, with substantial short-term fluctuation across the months. Although the death rate after 2019 remained elevated above the pre-2019 values, the upward trend became less pronounced with continued volatility marked by several spikes and dips.

Opioid and stimulant co-involved death rates had a generally upward trend over the years with a marked mean shift in 2020 and a peak (5.78 per 100,000) in 2021 (Supplementary file, Figure S1c). Following the mean shift in 2020, the upward trend was less pronounced with monthly fluctuations. Adjusting the level shift, sustained the trend and reduced the variance of the death rate values from the mean. All these time series showed a non-stationarity (Figure 1a – 1c), which was also evident through the gradually declining ACF plot and a PACF plot that dropped sharply (Supplementary file, Figure S1a – S1e). This was confirmed by an ADF test (p ≥0.05).

**Figure 1a:**
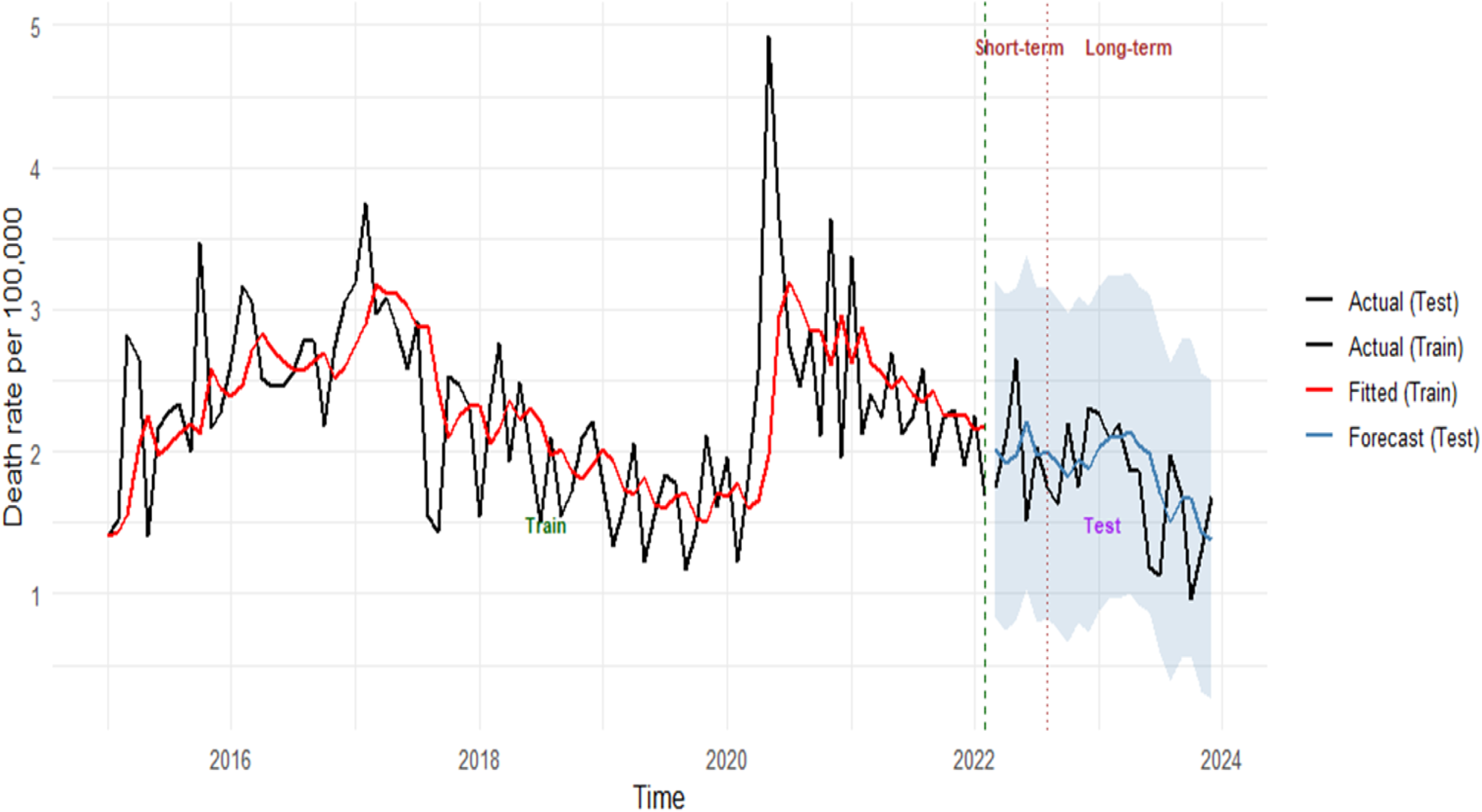
Autoregressive integrated moving average model for opioid-only death rates

**Figure 1b:**
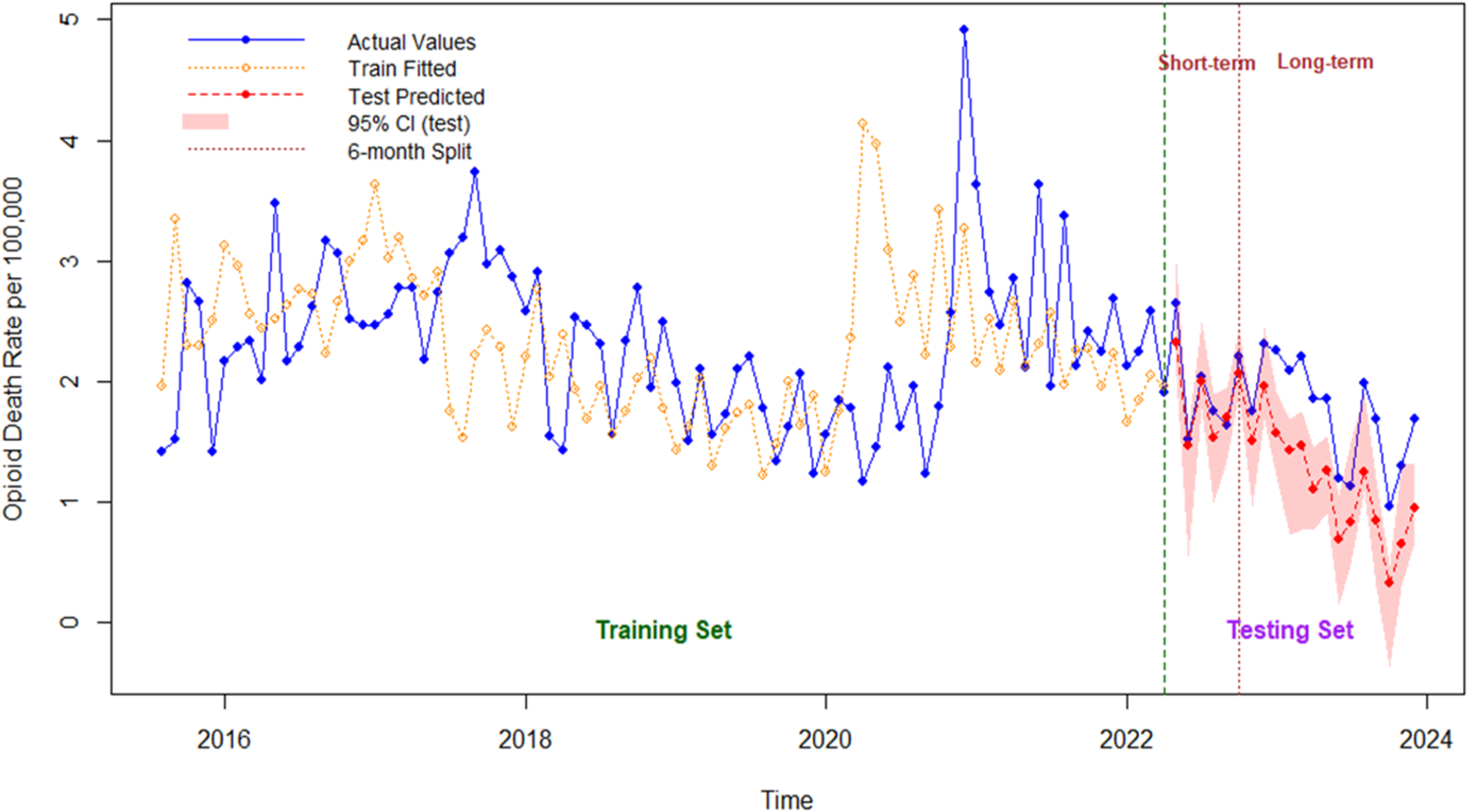
Random forest model prediction for opioid-only death rates

**Figure 1c:**
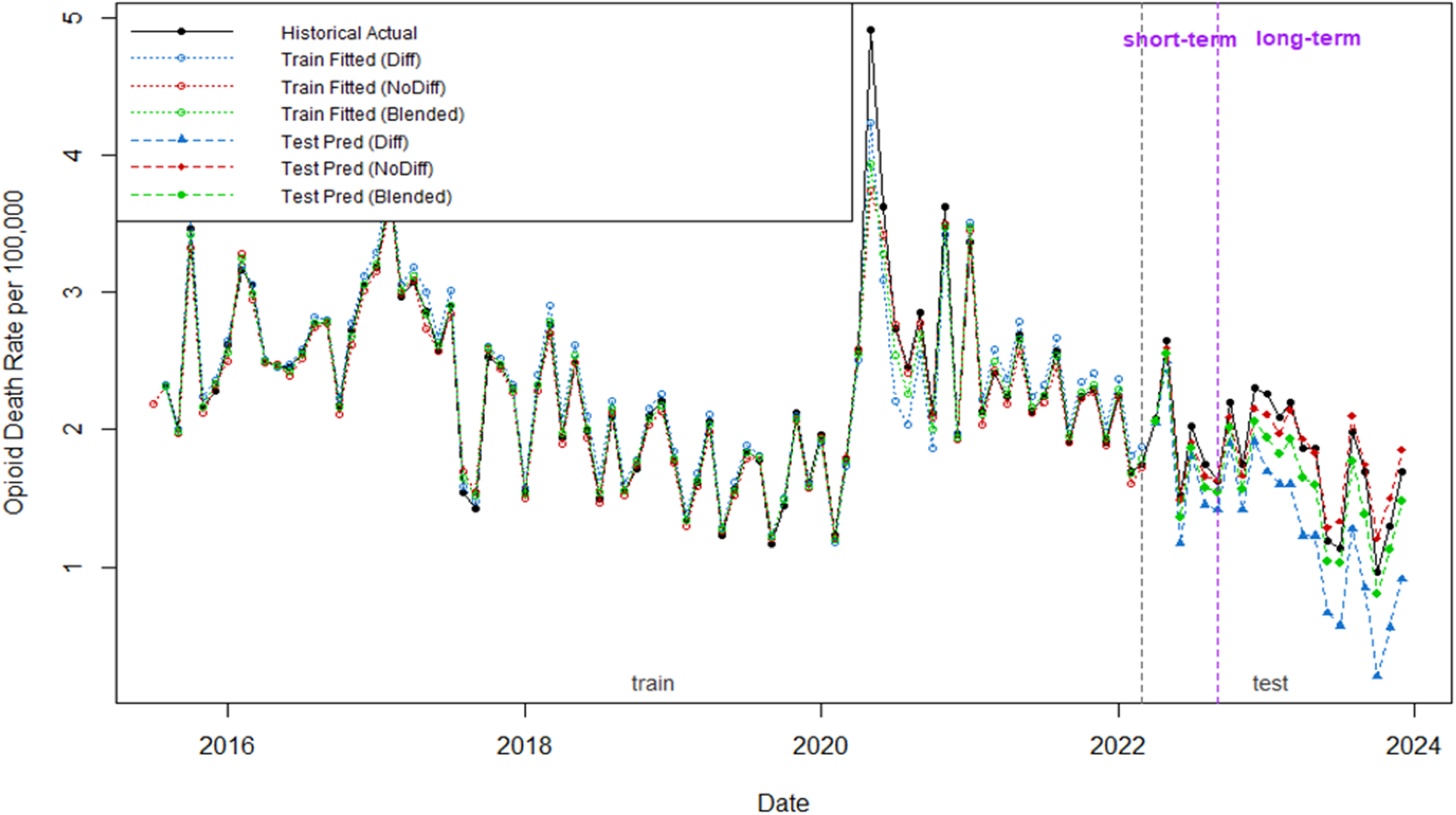
Differenced, non-differenced, and blended extreme gradient boosting model prediction for opioid-only death rates

### Model performances on Opioid-only Death Rates

Both opioid-only ARIMA models detected and predicted trends in the data but underperformed in capturing the definite peaks and drops in the data points (Figure 1a, Supplementary file, Figure S4a). No performance difference was observed between both models indicated by the test performance metrics (Table 2, R^2^ = 0.04, RMSE = 0.40, MAE = 0.33, MAPE = 21.00%). The models had a low predictive power (R^2^ = 0.04). All autoregressive coefficients of the models’ residuals were randomly distributed around the control line, which is equivalent to white noise and confirms the models’ validity (Supplementary file, Figure S5a – S5b). There was obvious variability between RF fitted and actual opioid-only death rate values in the train window (Figure 1b). The RF model predicted the trend in the test window with some alignment during the first 6 months but underestimated the values long-term. The model performed poorly (Table 2, R^2^ = −0.59), with a moderate average deviation between actual and predicted values (RMSE = 0.54, MAE = 0.47), and a 27.69% prediction difference from actual values. The non-differenced XGBoost model had the highest predictive power (Table 2, R^2^ = 0.92), more closely aligned test predictions for opioid-only death rate (Figure 1c), and the lowest error metrics (RMSE = 0.12, MAE = 0.10, MAPE = 6.59%).

**Table 2:** Model performance metrics for test predictions.

| Model | R<sup>2</sup> | RMSE | MAE | MAPE (%) |
| --- | --- | --- | --- | --- |
| **Opioid Death Rates** |  |  |  |  |
| ARIMA (actual values) | 0.04 | 0.40 | 0.33 | 21.00 |
| ARIMA (with imputed spike) | 0.04 | 0.40 | 0.33 | 21.00 |
| Random Forest | -0.59 | 0.54 | 0.47 | 27.69 |
| XGBoost (Differenced) | -0.63 | 0.53 | 0.48 | 29.93 |
| XGBoost (Non-differenced) | 0.92 | 0.12 | 0.10 | 6.59 |
| XGBoost (Blended) | 0.77 | 0.20 | 0.19 | 10.79 |
| **Stimulant Death Rates** |  |  |  |  |
| ARIMA | -0.24 | 0.24 | 0.19 | 27.70 |
| Random Forest | 0.22 | 0.20 | 0.16 | 22.94 |
| XGBoost (Differenced) | 0.14 | 0.20 | 0.17 | 24.10 |
| XGBoost (Non-differenced) | 0.91 | 0.07 | 0.06 | 7.35 |
| XGBoost (Blended) | 0.81 | 0.10 | 0.08 | 11.45 |
| **Opioid and Stimulant Death Rates** |  |  |  |  |
| ARIMA (Actual values) | -0.06 | 0.67 | 0.57 | 18.80 |
| ARIMA (with imputed level shift) | -0.10 | 0.69 | 0.58 | 33.70 |
| Random Forest | 0.57 | 0.44 | 0.35 | 12.14 |
| XGBoost (Differenced) | 0.74 | 0.34 | 0.26 | 8.77 |
| XGBoost (Non-differenced) | 0.67 | 0.38 | 0.32 | 10.16 |
| XGBoost (Blended) | 0.78 | 0.31 | 0.27 | 8.87 |

RMSE: Root mean square error, MAE: mean absolute error, MAPE: mean absolute percentage error

### Model Performances on Stimulant-only Death Rates

The ARIMA (0,1,1) model captured the overall trend in the data but underperformed at making point predictions of the values (Figure 2a). Model predictive ability was low (R^2^ = −0.24), although error metrics were not exceedingly high (Table 2, RMSE = 0.24, MAE = 0.19, MAPE = 27.70%). Residuals were randomly distributed (Supplementary file, Figure S5c). Visually, the RF model often overestimated peaks and underestimated drops in stimulant-only death rates during training (Figure 2b), although it was able to capture the trends in the test period. The model performed poorly with a low RMSE (0.22) but the deviation (Table 2, MAPE = 22.94%) between actual and predicted values was low. The non-differenced XGBoost model had the highest predictive power (R^2^ = 0.91), with the most closely aligned fitted and predicted values (Figure 2c), and the least variation between actual and predicted values (RMSE = 0.07).

**Figure 2a:**
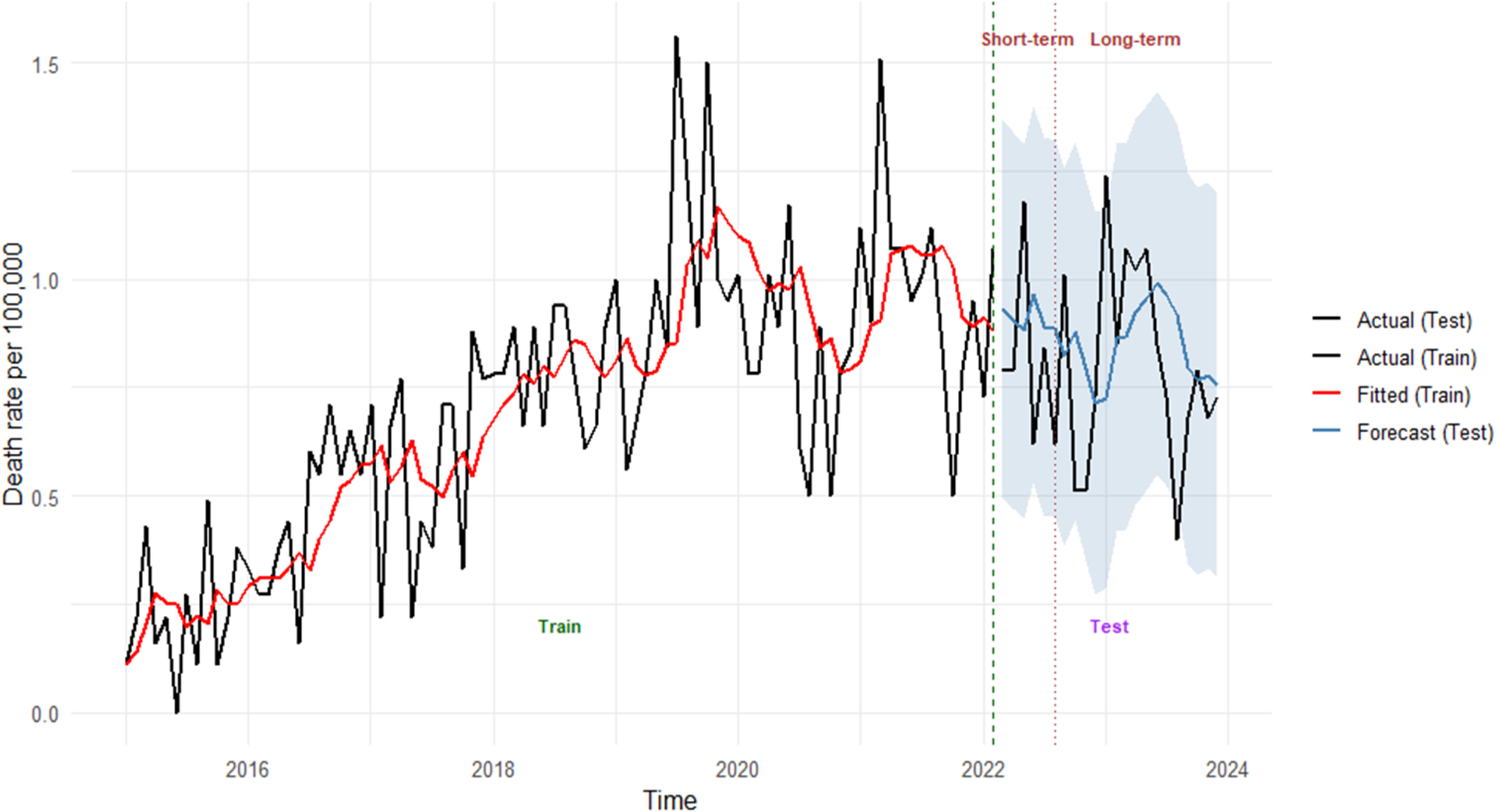
Autoregressive integrated moving average model for stimulant-only death rates

**Figure 2b:**
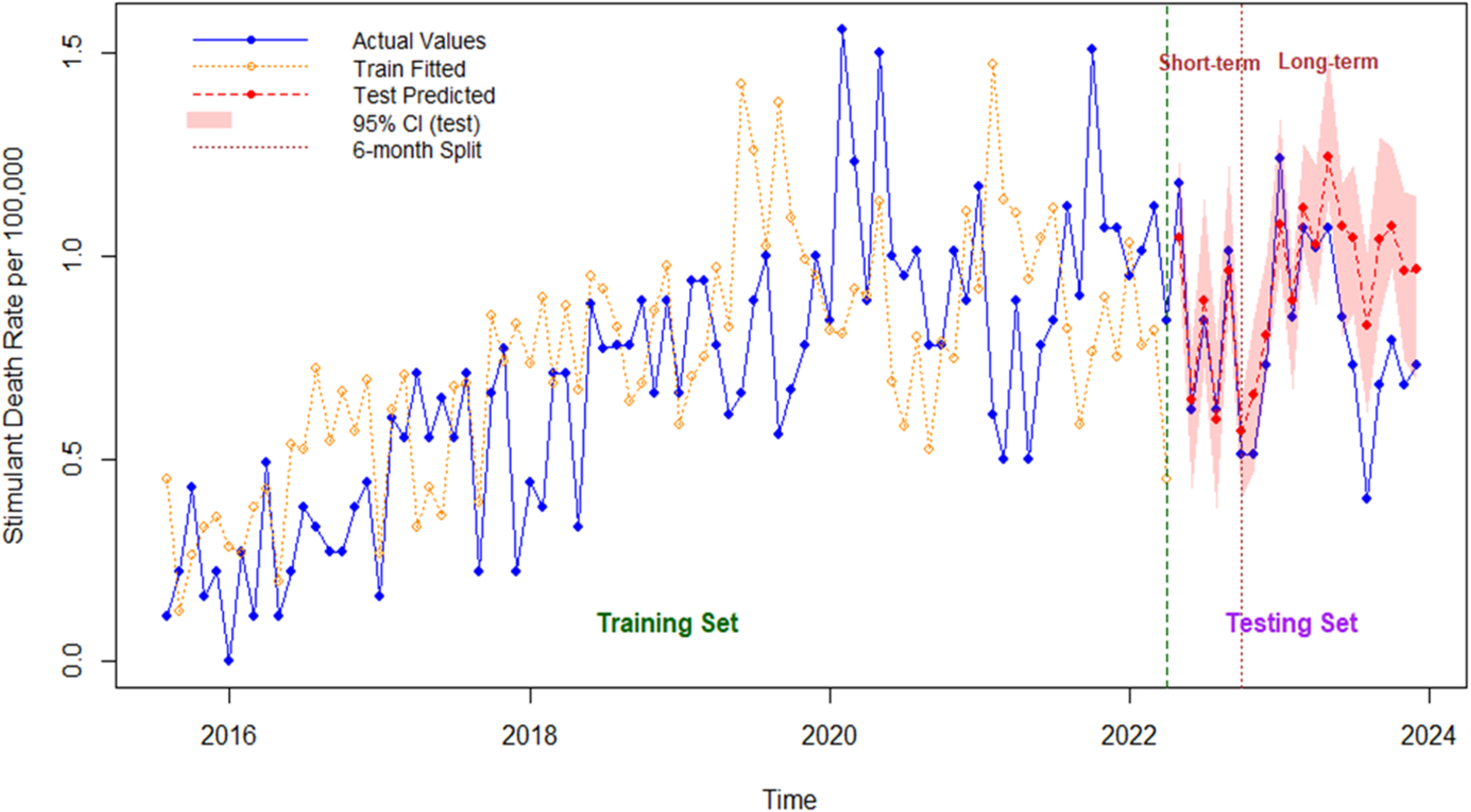
Random forest model prediction for stimulant-only death rates

**Figure 2c:**
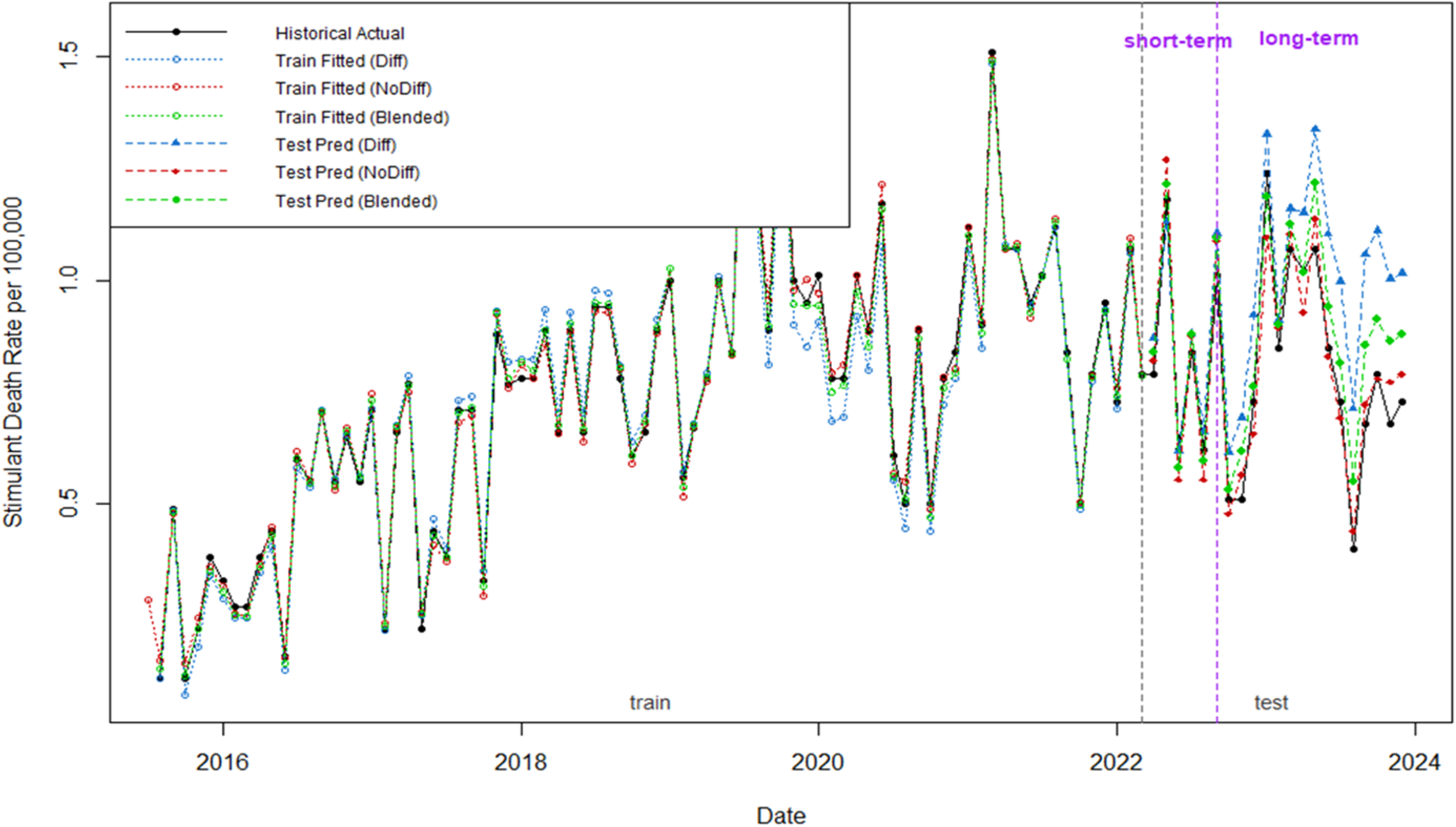
Differenced, non-differenced, and blended extreme gradient boosting model prediction for stimulant-only death rates

### Model Performances on Opioid and Stimulant Co-involved Death Rates

The ARIMA models captured the trend but there was visible variation between the fitted, forecasted, and actual values (Figure 3a, Supplementary file, Figure S4b). Although both models exhibited weak predictive power, (Table 2, R2 = −0.06 and −0.10), prediction error (RMSE = 0.67, MAE = 0.57, MAPE = 18.80%) were consistently lower for the model trained on the non-imputed death rate values. All residuals were randomly distributed (Supplementary file, Figure S5d – S5e). Although some values were overestimated, the RF model appeared to learn and replicate the trends in opioid and stimulant co-involved death rates (Figure 3b) with a moderate predictive power (R^2^ = 0.57) and a relatively minimal gap (MAPE = 12.14%) between the predicted and actual values. The non-differenced XGBoost model fitted the train data more closely (Figure 3c), but the blended model had the highest R^2^ value (0.78) and the least variation (RMSE = 0.31) between actual and predicted values for the test predictions.

**Figure 3a:**
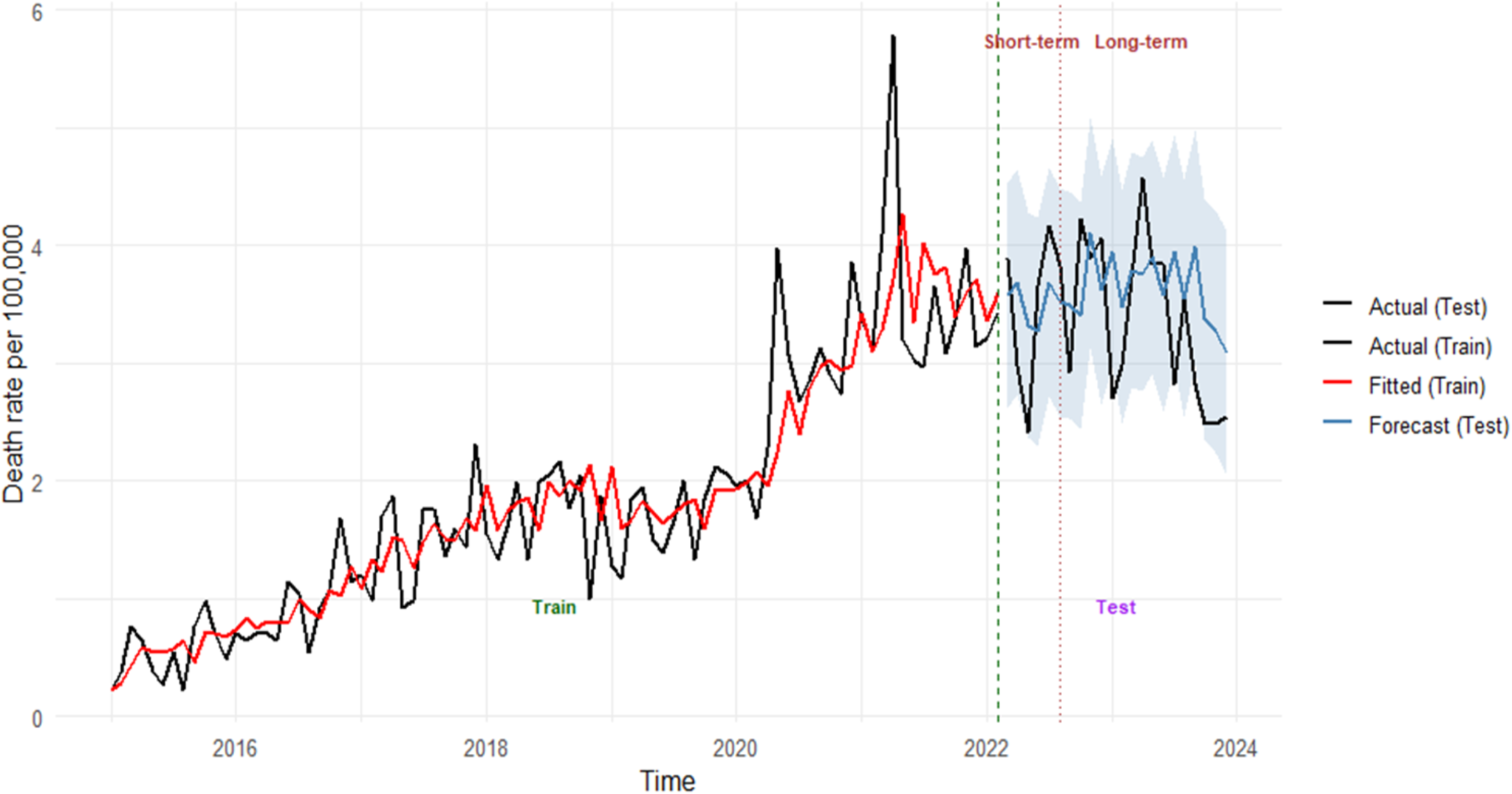
Autoregressive integrated moving average model for opioid and stimulant co-involved death rates

**Figure 3b:**
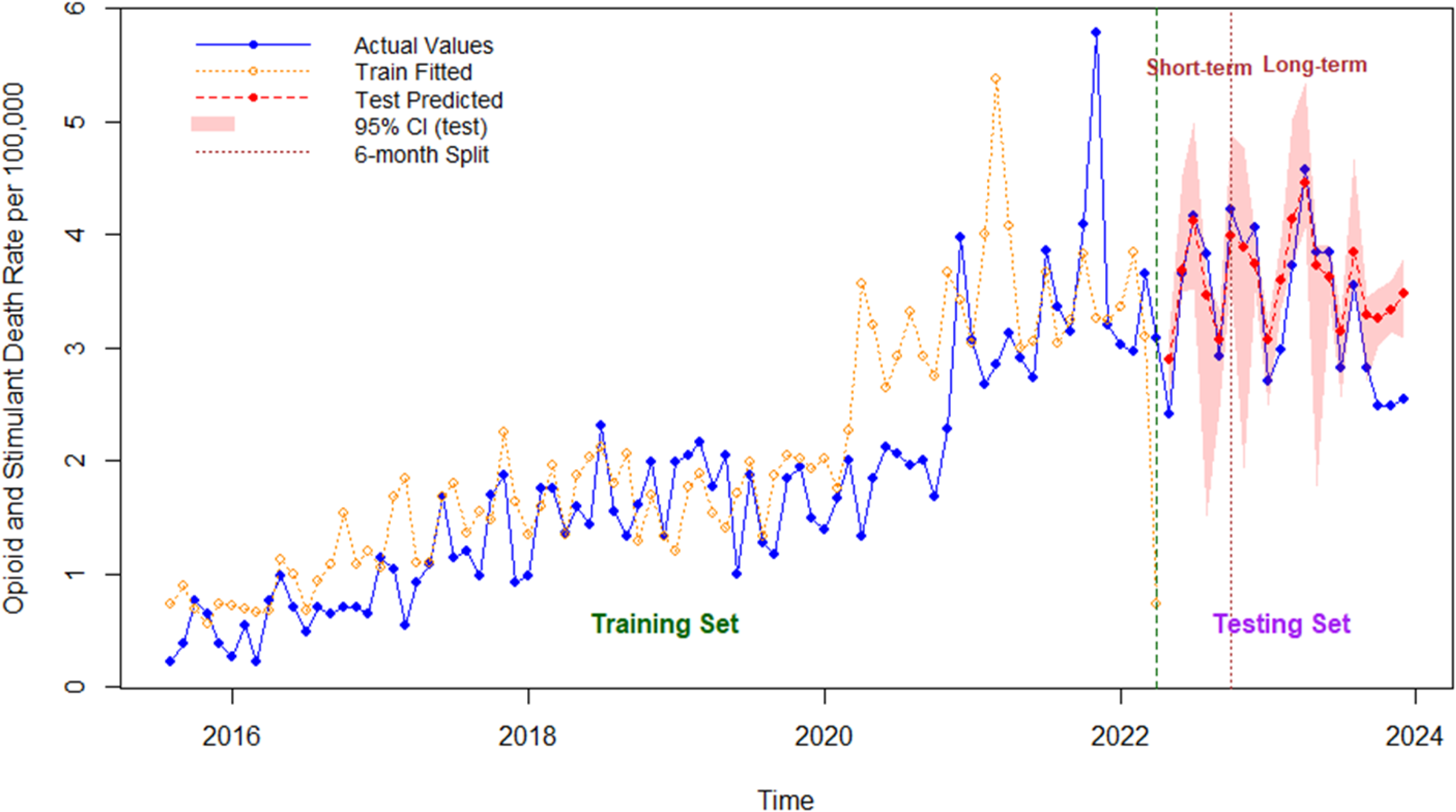
Random forest model prediction for opioid and stimulant co-involved death rates

**Figure 3c:**
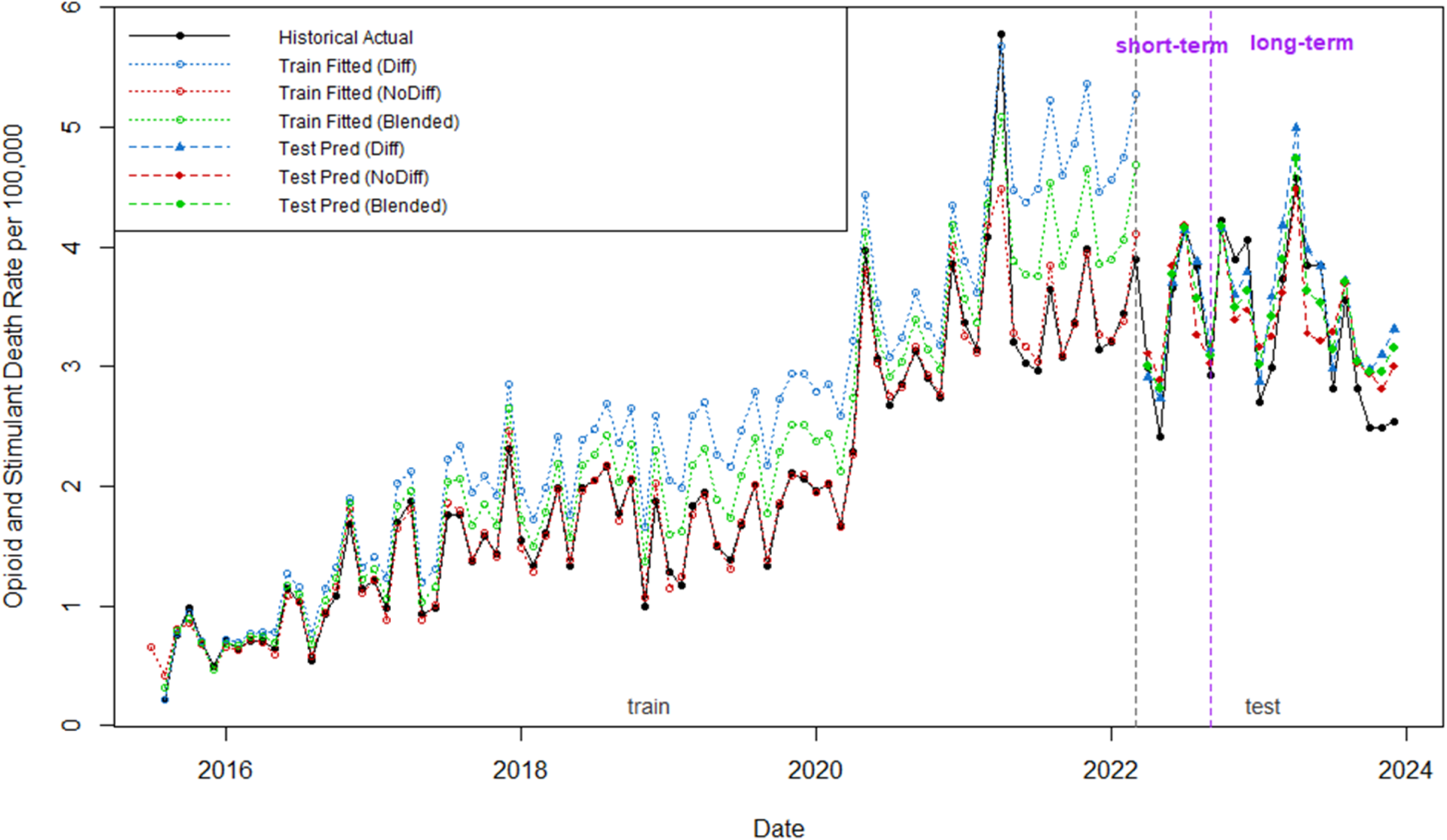
Differenced, non-differenced, and blended extreme gradient boosting model prediction for opioid and stimulant co-involved death rates

### Model Comparison

The non-differenced XGBoost model (R^2^ = 0.92) had better predictive power than ARIMA and RF in predicting opioid-only death rates, with the lowest error values (Table 2). Similarly, the non-differenced XGBoost model performed better than ARIMA and RF models for predicting stimulant-only death rates (R^2^ = 0.91, RMSE = 0.07, MAE = 0.06, MAPE = 7.35%). For opioid and stimulant co-involved death rates, the blended XGBoost model had the highest predictive power (R^2^ = 0.78) and operated with lesser error (RMSE = 0.31) compared to other models.

## Discussion

Concerns over rising opioid and stimulant overdose-related fatalities in the U.S. have been complicated by simultaneous misuse and abuse of both substances over time (2,25). Co-use of opioids and stimulants have been associated with more than twice the risk of fatal overdose compared to using opioids only (26). With the highest drug overdose mortality rate (81.9 per 100,000 persons) in the country in 2023 (27), WV also struggles with high rates of opioid and stimulant-related overdose deaths (3). Thus, there is dire need for high accuracy and precision surveillance and decision/policy support tools for predicting overdose fatality before its occurrence. Consequently, the predictions made can guide design and scaling of harm reduction interventions across the state, allocating resources and preparing health systems for overdose responses, and evaluating the impact of statewide efforts to control drug overdose mortality. Our findings revealed a continuous rise in WV monthly opioid and stimulant co-involved death rates with a peak in 2021. Stimulant-only death rate was highly variable, although the overall trend showed a continuous rise in fatality rate. In contrast, opioid-only death rates experienced a continual reduction except for a spike in 2020 followed by a gradual decline in fatality rate. Our comparative modeling of forecasting techniques to determine a suitable and best model for overdose death forecasting demonstrated relatively beneficial applicability of the ARIMA and RF models for predicting monthly overdose deaths involving opioids and stimulants alone or concomitantly. However, the XGBoost models compared to the other models, showed potential for even more accurate predictions with the non-differenced model performing best for opioid-only (R^2^ = 0.92) and stimulant-only (R^2^ = 0.91) death rates, while the blended model showed superiority for opioid and stimulant co-involved death rates forecasts (R^2^ = 0.78). These models exhibited comparatively better performance as a useful technique for drug overdose death surveillance and decision-making support.

The ARIMA model has an inherent ability to capture time dependence, the ability to model autocorrelational and seasonality components, and perform optimally with small datasets (12,28). Conversely, ARIMA struggles with capturing complex non-linear relationships, possesses limited built-in regularization ability to limit variance in predictions, and mostly performs best for short-term forecasts of univariate time series with clear patterns (29). In the opioid-only cohort, the ARIMA model underperformed compared to the non-differenced and blended XGBoost models which might be attributable to the relatively weak autocorrelation depicted by a rapid decay of the lagged values in the ACF plot (supplementary file, Figures 2a-2b). Other factors impacting the ARIMA models’ performances may be related to the absence of strong seasonality, non-linear patterns in the data, and a limited built-in regularization technique.

RF models can incorporate an unlimited number of external variables to make predictions without statistical constraints (30). It is suitable for complex multivariate scenarios with non-linear relationships and non-parametric data with no distribution assumptions, is capable of handling errors, and performs optimally on large datasets (30,31). However, RF models are computationally intensive and performance can be impacted by sample size (32,33). Predictive performance of an RF model is often limited in datasets with a strong temporal order since RFs treat observations independently and thus can completely miss casual or correlational dependencies (34). Performance of RF across the cohorts in our study was highly variable. The R^2^ values of the RF models were poor or moderate at best showing reasonably low forecasting ability, although RMSE, MAE or MAPE values were not exceptionally high. The poor performance may have resulted from the randomness in building multiple trees without correcting errors in past trees, time independence of predictions, and the inability to capture a strong temporal order. The relatively small number of datapoints could also be implicated in the RF models’ poor performances.

The shortcomings of RF are often ameliorated in XGBoost. The XGBoost model possesses automatic hyperparameter tuning and advanced L1 and L2 regularization features to prevent overfitting, and it can make predictions with greater accuracy and predictive power (17). Although XGBoost can be data intensive, its sequential learning approach enables it to also work with small datasets (17). In this study, these traits could account for the better performance of the non-differenced XGBoost model across the three cohorts. Developing different XGBoost models per cohort also introduced some interesting application perspectives. The differenced model makes short-term patterns more consistent and easier to model and the model responds quickly to recent shifts in the data, capturing immediate trends and fluctuations. The differenced model also reduced the impact of level shifts. Since differencing removes non-stationarity, the model has improved short-term accuracy and is suitable for short-term risk-assessment and predictions. However, it tends to accumulate error when reverted to the original scale, is highly volatile, and can miss persistent long-term trends. In our study, the differenced model captured sudden fluctuations in the stimulant-only death rates (Figure 2c), but often overestimated the values.

The non-differenced model holds the original scale and trend information and could capture slow-moving, persistent trends making it suitable for long-term predictions. Error compounding is avoided, and the forecasts are more stable with narrower confidence intervals less prone to explosive growth. The model exceled at predicting the actual level of the series instead of mere directional changes. Considering the stimulant-only cohort, the non-differenced model had superior performance and the visualization (Figure 2c) showed that the forecast tracked the actual values more accurately. Thus, we found the non-differenced model would be preferred for long-term forecasting and planning when there is a relatively consistent pattern over time and when identifying the overall underlying trend is preferred rather than focusing on short-term fluctuations. The blended model features a short-term responsiveness of differencing and the long-term stability of non-differencing which aids in mitigating extreme predictions and volatility. The fitted and forecasted values of the blended XGBoost model for opioid and stimulant co-involved deaths often fell between the differenced and non-differenced models. Hence, the blended model functions with a balance between short-term responsiveness and long-term stability to optimize forecasts. Consequently, the model will be most useful for making highly responsive yet stable overdose death forecasts.

Our findings build on other analyses of the relative performance and usefulness of ARIMA, RF, and XGBoost models in predicting public health outcomes, but applied here to model drug overdose mortality rates. The machine learning models utilized in this study introduced innovative predictive approaches, expanding the methodological toolkit for drug overdose death surveillance. The ability to predict future increases in drug overdose fatalities with greater accuracy can provide overdose death rate estimates that can be referenced during planning and allocation of resources for overdose death prevention. Our work can enhance the accuracy and reliability of overdose fatality rate forecasts, especially when traditional statistical methods fall short.

To the best of our knowledge, this is the first study to employ both statistical and machine learning models to forecast single and polysubstance-related overdose fatality rates using real-world data especially one using data from a specific state. Our comparative evaluation offers a robust view of short-and long-term forecasting capabilities for overdose deaths and highlights the strengths and limitations of each model. Our study also uncovers any unique features related to each time series and provide a scalable group of models to handle different natured time series data. For example, opioids and stimulants may exhibit varying temporal patterns when implicated individually or simultaneously in overdose fatality, with one depicting more seasonal variation and trend consistency than the others. Three different XGBoost models were developed in this study that can be applied for short-term surveillance of overdose fatalities or long-term forecasting for planning and resource allocation purposes. The purpose of the forecasting, whether for short-term trend monitoring, long-term predictions for planning purposes or both, will determine the appropriate model to use. Importantly, the machine learning models can be updated and retrained on new data, adapting the approach to changing overdose death patterns.

Database size poses a considerable limitation to the performance of the developed models. RF and XGBoost are data intensive and perform optimally with larger datasets. However, our data had 108 data points which may have been insufficient to learn temporal patterns in the time series data. Although ARIMA can perform relatively well with few datapoints, more data are often required to detect and learn detailed seasonal and temporal patterns. Therefore, the models’ performances may have been limited by the data size. Unlike ARIMA, RF and XGBoost are often more difficult to interpret, possibly limiting their transparency for public health decision-makers. The overdose deaths trends had a non-linear pattern which deviated from the linearity assumption of the ARIMA model, which probably impacted the performance of ARIMA in this study. Since the models were trained on WV data from a specific timeline, they may not generalizable adequately to other settings and time frames without retraining and calibration. The forecasts only identify patterns but do not establish causal relationship, which restricts their use for identifying the root causes of changes in overdose death rates for specific substances.

In conclusion, machine learning models such non-differenced XGBoost model performed best for predicting single substance (opioid-only and stimulant-only) death rates. The blended XGBoost model had the best predictive performance for polysubstance (opioid and stimulant co-involved) death rates. Also, XGBoost model showed superiority on both short- and long-term forecasting over other models. These models can be valuable for forecasting and monitoring drug overdose deaths although the optimal model to use might vary based on the attributes of the substances involved and whether short-term or long-term predictions are desired. Future studies should examine the role of opioid and stimulant prescription rates as predictors of their related fatality rates while incorporating more years of data and integrating the role of illegal drugs. These models can also be applied to investigate trends for newly emerging substances of abuse such as xylazine and other substance analogs.

## Materials and Methods

### Data Acquisition and Preparation

West Virginia drug overdose death data from 2015 to 2023 were obtained from the forensic research data (FRD). Data was assessed from January 01, 2015, to December 31, 2023. The FRD was developed through a collaboration with the West Virginia Office of the Chief Medical Examiner (WVOCME) to compile drug-related death information in the state (9). Case entries in the FRD contain demographic information (e.g., age, sex, county, and zip code of residence), date of death, manner of death, identified drugs, drug involvement, drug count, and whether prescriptions were present for controlled substances involved in the death (9). Information in the FDD is compiled from multiple sources, which include death certificates, autopsy reports, medical records, external examination, investigator reports, police reports, toxicology reports, and the WV Controlled Substances Monitoring Program (WVCSMP) (9). Since the individual records are de-identified, this study was deemed exempt from Institutional Review Board (IRB) approval by the West Virginia University IRB.

All opioid and stimulant-involved deaths were included. Non-opioid and non-stimulant-associated cases were excluded. From the identified cases, three different cohorts were formed: opioid-only, stimulant-only, and opioid and stimulant co-involved fatalities. The opioid-only cohort included decedents with any opioid (see Supplementary file, Table S1) and no stimulant involved death, the stimulant-only cohort included those with any stimulant and no opioid involved death, and the opioid and stimulant co-involved death cohort included decedents who had at least one opioid and one stimulant involved in their death. For each cohort, the number of cases in each month for a particular year was obtained, and the monthly death rates per 100,000 were calculated, with the number of cases per month divided by West Virginia’s population for that year (10,11), then multiplied by 100,000. A time series plot with 108 data points was generated using the death rates of each cohort.

### Autoregressive Integrated Moving Average Model

Autoregressive Integrated Moving Average Model (ARIMA) is a statistical model with a time-dependent structure, designed for time series analyses, leveraging its built-in components for detecting trend and seasonal data patterns (12). The ARIMA model is depicted as ARIMA (p,d,q), where p is the order of autoregression, d is the degree of differencing, and q is the moving average term (13).

ARIMA requires stationarity of the time series data. An Augmented Dickey Fuller (ADF) test was conducted on the monthly death rate data for each cohort to test for non-stationarity. A p-value > 0.05 confirmed non-stationarity for each cohort, with first-order differencing applied to the death rates as appropriate to make them stationary. A repeat ADF test on the differenced data confirmed stationarity (p < 0.05). Each cohort’s data was split into 80% training and 20% test sets with sustained time ordering. The p and q terms for the ARIMA models were determined based on the partial autocorrelation function (PACF) and autocorrelation function (ACF) plots (Supplementary file, Figure S3a – S3e), respectively, of the differenced death rates for the respective cohorts. The ACF plots the dependence of a specific observation of a variable and its previous values at different time points (14). PACF measures the direct correlation between an observation and past values without the influence of the correlation of intermediate values (14). To ensure appropriate model selection, different ARIMA models (ARIMA (0,1,1), (1,1,1), & (2,1,1)) were compared for each cohort, and the model with the least Akaike Information Criterion (AIC) was chosen. Thus, ARIMA (0,1,1) was selected for opioid-only and stimulant-only, while ARIMA (2,1,1) was selected for the opioid and stimulant co-involved death cohort. An ARIMA model was trained on the training set and then utilized to predict the test data. A residual test was conducted to assess if the model adequately captured all relevant information from the time series, confirmed by a homogenous pattern in the residual plot. To account for the impact of spike/pulse or level shift on the fit and forecasting ability of an ARIMA model, two distinct ARIMA models were each developed for opioid-only and opioid and stimulant co-involved death rates. The first model utilized the calculated death rate data for each cohort. In the second model, the opioid death rate spike in 2020 was imputed with the mean of the values before and after the spike. The level shift in opioid and stimulant co-involved death rate (2020–2023) was imputed by subtracting the difference of the mean before and after the shift from values within the level shift.

### Random Forest Model

The RF model is a machine learning ensemble technique that functions by constructing multiple, internally validated decision trees from a training dataset before aggregating them to make a prediction (15). Model feature engineering, including lags, rolling statistics, and momentum features, was conducted to represent temporal relationships necessary for forecasting. First-order differencing was done to address non-stationarity in the death rates of each cohort. Six temporal lag features (Lag1 – Lag6) were created from the death rate data of each cohort to enable capturing of direct autocorrelation data patterns. Three-and six-month rolling means were calculated to smooth short-term fluctuations to reveal quarterly trends and to capture medium-term trends spanning half-year periods, respectively. Momentum features, including first-order differencing (Differencing 1), deviation from 3-month average (Differencing 3), and percentage change relative to previous values (Differencing percent), were determined. The momentum features were necessary to measure immediate acceleration or deceleration in death rates, identify anomalies from recent trends, and highlight proportional shifts independent of absolute values, respectively. Calendar features (month, quarter, and year) were excluded because they reduced the models’ usefulness due to numerous possible predictors. Also, RF lacks L1 and L2 regularization for model selection. Next, the top five (5) features of the model were selected, including differencing 3, differencing 1, three-month rolling mean, six-month rolling mean, and lag1, based on node purity importance ─ a measure of the tree classification confidence (16). Afterward, the model was configured with 500 trees, node size = 5, and full variable importance tracking. The data for each cohort were split into an 80% training set and a 20% test set while preserving time ordering. A recursive multi-step forecasting where each prediction becomes an input for future steps was conducted to predict the test set values. The 95% confidence intervals were derived using the forestError package in R or standard error approximation.

### Extreme Gradient Boosting Model

XGBoost is a tree-based model with a sequential tree-building approach to minimize prediction errors (17). New trees are built while correcting for errors made by a combination of the previous trees (17). The XGBoost model was manually feature engineered for time series forecasting to enable it to learn complex temporal patterns through sequential optimization.

Three different XGBoost models (differenced, non-differenced, and blended) were developed for each cohort. In the blended model, a weighted ensemble containing 40% of the differenced and 60% of the non-differenced models was created. Lag values, rolling means, and the momentum features were derived, and the top five features were selected, similar to the RF models. The data were split into an 80% training set and a 20% test set with time ordering. Bayesian optimization was applied to tune eta, max_depth, and min_child_weight parameters. Early stopping with 20-round patience was implemented to avoid overfitting. Recursive multi-step forecasting was also used to predict monthly death rates for the test duration. 95% prediction confidence intervals were generated via bootstrapping residuals for each model.

### Statistical Analysis and Model Comparison

R^2^, root mean square error (RMSE), mean absolute error (MAE), and mean absolute percentage error (MAPE) were computed for the test predictions for each ARIMA, RF, and XGBoost model. Model performance on test predictions for each cohort was compared using these parameters. The R^2^ value is critical in determining the extent of the total variation in the time series attributable to each model, with a higher value signifying a good model fit (18). On the other hand, RMSE estimates the magnitude of variation between actual values of a time series and the predicted values by a model; hence, lower values are preferred (18). Additionally, MAE presents the average of the absolute values of forecast error, while MAPE measures how much a time series differs from its model-predicted values as a percentage (18).

All data preparation steps were conducted using R programming version 4.4.0. The forecasting steps were conducted on R programming using the tseries, forecast, xgboost, randomForest, forestError, and parBayesianOptimization packages (19–24).

## Acknowledgements

Not applicable

## Data availability statement

The deidentified decedents data supporting this study was obtained from the West Virginia Office of the Chief Medical Examiner and not available for public sharing due to its sensitive nature.

## Supplementary files

Table S1: Drug names

Figure S1a: Time series plot for opioid-only death rates from 2015 – 2023

FigureS1b: Time series plot for stimulant-only death rates from 2015 -2023

Figure S1c: Time series plot for opioid and stimulant co-involved death rates from 2015 – 2023

Figure S2a – Autocorrelation function and partial autocorrelation function plots of opioid-only death rate values

Figure S2b – Autocorrelation function and partial autocorrelation function plots of opioid-only death rate values with imputed spike

Figure S2c – Autocorrelation function and partial autocorrelation function plots of stimulant-only death rate values

Figure S2d – Autocorrelation function and partial autocorrelation function plots of opioid and stimulant co-involved death rate values

Figure S2e – Autocorrelation function and partial autocorrelation function plots of opioid and stimulant co-involved death rate values with imputed level shift

Figure S3a: Autocorrelation function and partial autocorrelation function plots of differenced opioid-only death rate values

Figure S3b: Autocorrelation function and partial autocorrelation function plots of differenced opioid-only death rate values with imputed spike

Figure S3c: Autocorrelation function and partial autocorrelation function plots of differenced stimulant-only death rate values

Figure S3d: Autocorrelation function and partial autocorrelation function plots of differenced opioid and stimulant co-involved death rate values

Figure S3e: Autocorrelation function and partial autocorrelation function plots of differenced opioid and stimulant co-involved death rate values with imputed level shift

Figure S4a: Autoregressive integrated moving average model for opioid-only death rates with imputed spike value

Figure S4b: Autoregressive integrated moving average model for opioid and stimulant co-involved death rates with imputed level shift

Figure S5a: Residual plot of autoregressive integrated moving average model for opioid-only death rate

Figure S5b: Residual plot of autoregressive integrated moving average model for opioid-only death rate with imputed spike

Figure S5c: Residual plot of autoregressive integrated moving average model for stimulant-only death rate

Figure S5d: Residual plot of autoregressive integrated moving average model for opioid and stimulant co-involved death rate

Figure S5e: Residual plot of autoregressive integrated moving average model for opioid and stimulant co-involved death rate with imputed level shift

